# Longitudinal Associations of Dietary Fructose and Psychological Stress with Vascular Aging Index and Incident Cardiovascular Disease in the CARDIA Cohort

**DOI:** 10.1101/2023.10.03.23296515

**Authors:** Meaghan Osborne, Alexa Bernard, Emily Falkowski, Deni Peterson, Anusha Vavilikolanu, Dragana Komnenov

## Abstract

**Background:** Cardiovascular disease (CVD) risk increases exponentially with age, largely due to vascular aging. We explored how dietary behaviors (sucrose, fructose, sodium, and potassium consumption) and endured psychological stress in young adult males and females impact the vascular aging index (VAI) and CVD risk by mid-life.

**Methods:** Data were obtained from the Coronary Artery Risk Development in Young Adults Study, an ongoing longitudinal study. Included participants (n=2,656) had undergone carotid artery ultrasound Doppler scans at year 20 allowing VAIs to be calculated. Demographics, dietary data and depression scale scores were obtained at the initial visit. Regression analyses were used to assess the predictors of VAI 20 years later. Cox regression analyses were conducted to assess the risk of fatal and non-fatal CVD, hypertensive CVD, and stroke.

**Results:** Predictors of vascular aging were found to be sex-specific. In females, CES-D scores at baseline were positively associated with VAI (B-weight=0.063, p=0.015). In males, sodium intake at year 20 positively predicted VAI (B-weight=0.145, p=0.003) and potassium intake inversely predicted VAI (B-weight=-0.160, p<0.001). On Cox regression analyses, BMI significantly predicted CVD, stroke, and death. In addition, fructose consumption at year 20 was a significant predictor of CVD risk while having blood pressure above 130/80 mmHg at baseline was significantly associated with stroke risk.

**Conclusion:** Our findings support the promotion of nutrient-specific behavior changes (specifically, limiting fructose consumption) to prevent vascular aging in early adulthood and CVD risk in mid-life.

## INTRODUCTION

Cardiovascular disease (CVD) is the leading cause of death and disability worldwide ^1^. CVD risk increases exponentially with age, largely due to pathophysiological changes in the vasculature, namely calcification, loss of elastin, increased collagen deposition, and increased vascular diameter^2^. The major consequence of vascular aging is increased arterial stiffness, which decreases compliance and the ability of the vessel to adapt to stressors ^2^. The gold standard measurement of arterial stiffness is carotid-femoral pulse-wave velocity (cfPWV) which has been shown to be a strong predictor of future CVD events ^3^. Recent data suggests that the combination of the vessel structural property (i.e. carotid intima–media thickness (cIMT)) with the functional property (i.e. PWV) into a vascular aging index (VAI) may improve the prediction of CVD risk ^4^. Many mechanisms have been proposed to explain the vascular aging process including oxidative stress, chronic inflammation, and the cellular mechanisms driven by mammalian target of rapamycin signaling pathway ^5^. In addition, it is well known that hypertension leads to early vascular aging ^6^, as well as certain lifestyle factors including psychological stress and diet ^7-9^. Given that the U.S. population over 65 years is expected to nearly double by 2060, and that age is a major risk factor for CVD, there is a need to elucidate the mechanisms that affect vascular aging process ^10^.

According to the 2018 National Health Interview Survey, males have a higher prevalence of heart disease, coronary heart disease, hypertension, and stroke (12.6%, 7.4%, 26.1%, and 3.1%) when compared to age-matched females (10.1%, 4.1%, 23.5%, and 2.6)^1^. Sex hormones, mainly estrogens, have been used to explain such differences, given that the post-menopausal period is associated with increased CVD prevalence and heart failure development in females^1,11^. One of the mechanisms proposed includes estrogen-mediated β-adrenergic vasodilation ^12^. The hypothesis that estrogen is cardioprotective is supported by the finding that pre-menopausal arteries have significantly less endothelial inflammation in the setting of enhanced vasodilatory capacities^13^. This in turn prevents oxidative damage, which is a significant mediator of arterial stiffening and thus vascular aging ^14^. Any protective effects of estrogen pre-menopause appear to be eliminated post-menopause whereby post-menopausal females experience a rapid increase in blood pressure, calcification scores, and central arteriole stiffness ^14^.

In addition to intrinsic phenotypic sex differences, it is now recognized that extrinsic or psychosocial factors also contribute to the sex differences in CVD risk and vascular aging ^14^. For example, females are at a two-fold greater risk of experiencing depression compared to males and depression is a known CVD risk factor ^15^. Adolescent females may experience more interpersonal stress ^16^. In addition, childcare responsibilities may impact the prevalence of depression and anxiety in females more than males ^17^. The effects of depression on vascular aging have been largely unexplored. Given the sex and gender differences in CVD risk factors, CVD outcomes, and vascular aging, it has been suggested that future studies should incorporate sex disaggregated data for traditional and non-traditional risk factors to help inform precision medicine ^18^.

Diet constitutes a modifiable lifestyle factor with potential consequences for vascular aging. According to the AHA’s Life’s Essential 8, a diet rich in fruit and vegetables, complex carbohydrates, fish, beans, fiber, moderate in meat, and minimal in processed sugary foods can prevent endothelial dysfunction and arterial stiffness ^7, 19^. While these recommendations offer broad, healthy dietary patterns, there has been an impetus for studies focusing on specific micro- and macronutrients within those patterns for promoting healthy vascular aging ^20^. One relatively well-studied dietary component is sodium. Reducing sodium intake has been shown to consistently lower arterial stiffness in randomized controlled and crossover trials with normotensive and hypertensive middle-to-older aged males and females ^21^. Limitations of these clinical trials include the small sample sizes and limited racial and ethnic diversity ^22^. Moreover, a recent trial in individuals with heart failure (HF) showed that reducing dietary sodium did not improve clinical events compared to those HF patients who did not reduce dietary sodium ^23^. Indeed, additional epidemiological data is needed exploring the associations between dietary sodium and vascular aging in the context of covariates important for CVD development: sex, race and ethnicity, hypertension status, and other dietary culprits, such as the high fructose corn syrup (HFCS). Given that dietary potassium may also play a vital role in modulating CVD risk, exploring the effects of potassium intake on vascular aging is also warranted ^24, 25^.

Dietary sugar is a disaccharide of glucose and fructose, but an important distinction exists in the fructose component in North American countries compared to some of the European countries (i.e. Scandinavia). Where sugar-sweetened processed foods contain HFCS, the fructose component is typically in excess to glucose by about 50% ^26, 27^. In fact, since the advent of HFCS in the 1970’s, this has been the primary component of sugar used by the food industry in North America ^28^. Indeed, pre-clinical work showed that even short -term moderate increase in dietary fructose causes salt-sensitive blood pressure, diastolic dysfunction and increased aortic PWV in rats ^29-32^. Additionally, other rodent models showed that increased ingestion of fructose and salt during early age (i.e. equivalent to adolescence in humans), contributes to hypertension, vascular stiffness and decrease in glomerular filtration rate (GFR) later in life even after the exposure to fructose and salt had been removed and later re-introduced to rats once they were older (i.e. equivalent to mid-life in humans) ^33, 34^. Likewise, in a systematic review and meta-analysis of six human cohort studies, sugar-sweetened beverages were significantly associated with an increased risk of hypertension, although vascular aging variables were not measured ^35^.

In the present investigation we included the participants from the CARDIA (Coronary Artery Risk Development in Young Adults) study. To our knowledge, few studies have explored the role of diet on vascular aging within the CARDIA cohort. Gao et al ^36^ found that among CARDIA participants, carbohydrate intake was negatively correlated with the risk of coronary artery calcium progression, a marker of atherosclerosis. Duffey et al ^37^ showed that sugar-sweetened beverage consumption was positively associated with waist circumference, triglycerides, LDL cholesterol, and hypertension. The objective of the present retrospective observational study was to assess how dietary behaviors, specifically fructose and sodium, and endured psychological stress in young adult males and females impact the VAI and CVD risk by mid-life. As the burden of age-associated CVD continues to climb, the need for novel preventive health approaches to vascular aging is paramount.

## MATERIALS AND METHODS

### Study Sample

The study sample included the participants from the CARDIA study designed to examine lifestyle and behavioral factors that contribute to the development of CVD from young adulthood through midlife. The study recruited 5,114 individuals, aged between 18 and 30 years (median = 26 years) in 1985 and 1986 at four centers across the United States: Birmingham (AL), Chicago (IL), Minneapolis ^33^ and Oakland (CA), matched for sex, age, race (Blacks and Whites) and education level. Assessments occurred at baseline and at years 2, 5, 7, 10, 15, 20, 25, and 30 (nine total assessments to date). The details on the study were published elsewhere ^38^. Here we considered the records from N = 2,656 participants for which carotid ultrasound scans were completed and carotid PWV obtained during visit 7 (year 20 of follow-up). The baseline values from visit 1 and outcomes (CVD, hypertensive CVD and stroke) from the latest visit (follow-up year 35) were obtained only for these individuals (N = 2,565) rather than an entire starting cohort of 5,114 people. Therefore, no missing data were observed for the cPWV and cIMT which were used in the derivation of VAI. Overall missing data were less than 5% for the remaining variables, and they were not imputed. The study was approved by the Institutional Review Board at Wayne State University (IRB # 063319MP2X). Most recent approval of the Research Materials Distribution Agreement (RMDA) by the National Institutes of Health, Heart, Lung and Blood Institute was completed on April 15, 2022.

### Study Measures and Outcomes

VAI was obtained as described previously ^4^, with modification of using carotid PWV (cPWV) instead of aortic PWV (aPWV). The value for cPWV was calculated by averaging the PWV values obtained on the left and right-side vessels expressed in the units of m/s. Carotid IMT (cIMT) was expressed in mm. The modified equation used to calculate VAI was:

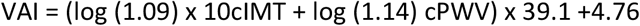

Blood pressures were measured in triplicates with a random zero mercury sphygmomanometer for visit 1 by centrally trained personnel. For visit 7, an Omron HEM907XL oscillometric monitor was used for obtaining BP values in triplicates after a 5 min rest period using the participants’ right arm. The inter-device differences were corrected by calibration as described before ^39^. Observed high blood pressure (obsHBP) was categorized for clinical elevation (>130 mmHg/80 mmHg, as per the American Heart Association criteria ^40^ at each time point. At baseline, 12.8 % of individuals (339 out of 2656) had obsHBP, independent of hypertension diagnosis, and in year 20, 6.4% (n = 171 of 2656) had obsHBP.

Dietary data were collected using the diet history method at visits 1, 4, and 7 (baseline, year 7, and year 20) referencing intake for the previous month. For average daily intake calculations (for sucrose, fructose, sodium and potassium) it was assumed that one month had 30.42 days. Nutrient amounts were calculated with Version 20 of the Food Table from the Nutrition Coordinating Center at the University of Minnesota. Regarding outliers and missing values, the Nutrition Working Group during visit 1 decided to include all out-of-range values in the participants’ records and to not impute any missing data on serving amounts or frequency. Instead of using absolute number of grams for sucrose and fructose, we transformed these values to percent total calories by multiplying the number of grams of sucrose/fructose by 4, dividing this number by the total number of calories and multiplying by 100.

Psychological stress was assessed using the Centre for Epidemiologic Studies Depression Scale (CES-D), a validated self-report psychometric scale. Items 1-20 are scored on a four-point Likert scale and then summed to obtain a total score that ranges from 0-60, whereby a higher score indicates more depressive symptomatology.

Physical activity levels were collected using a validated, self-reported questionnaire of leisure time and occupational physical activity ^41^. Participants were asked about their participation in moderate (e.g. walking, golfing, bowling) and vigorous-intensity (e.g. biking, running, racquet sports) aerobic activities over the previous 12-month period.

Our primary, second, and tertiary outcomes (CVD, stroke, and all-cause mortality, respectively) were obtained from the most recent visit at year 35 when the cohort was between the ages of 53 and 65.

### Statistical analysis

For statistical analysis, we used Statistical Package for the Social Sciences (SPSS) version 28.01.0. Prior to analysis, all continuous variable scales were screened for univariate normality with Kolmogorov-Smirnov test and most measures were found to have issues with skewness and kurtosis. A total of 2,656 individuals were included in the analysis. We used numbers and percentages for the participant’s characteristics description, and comparisons between males and females were made with a Pearson χ2 test. Race and sex were self-reported. Statistical significance was identified with *p*-values of <0.05. We completed multivariate linear regression analyses for factors associated with VAI in males and females. Collinearity of regression coefficients was ruled out with Pearson correlation and Spearman’s rho. We used Cox proportional hazards regression and corresponding hazards ratios for factors associated with primary, secondary and tertiary outcomes: development of any CVD (fatal and non-fatal), development of stroke (fatal and non-fatal), and death, respectively.

## RESULTS

A total of 2,656 patients were included in the final analysis. There were 1159 males (44%) and 1497 females (56%). Participant characteristics are presented in Table 1. The two groups were significantly different for dietary patterns, whereby males had higher sodium intake both at baseline and at year 20 of follow-up (p <0.001 for both) while females had higher sucrose intake at year 20 of follow-up (p = 0.002). Females had higher depression scores, expressed as Center for Epidemiological Studies Depression (CES-D; p = 0.021) at year 20 of follow-up. Males had lower left and right carotid PWV compared to females (p < 0.001 for both), while they had higher carotid IMT (p < 0.001). Consequently, VAI was found to be higher in males compared to females (p <0.001). High blood pressure diagnosis was more frequent in males than females at baseline, while the cohort was between 18 and 30 years of age, while this difference was no longer present 20 years later. At baseline, heart and kidney problems were more frequent in females compared to males (p = 0.007 and p < 0.001, respectively).

**Table 1.**
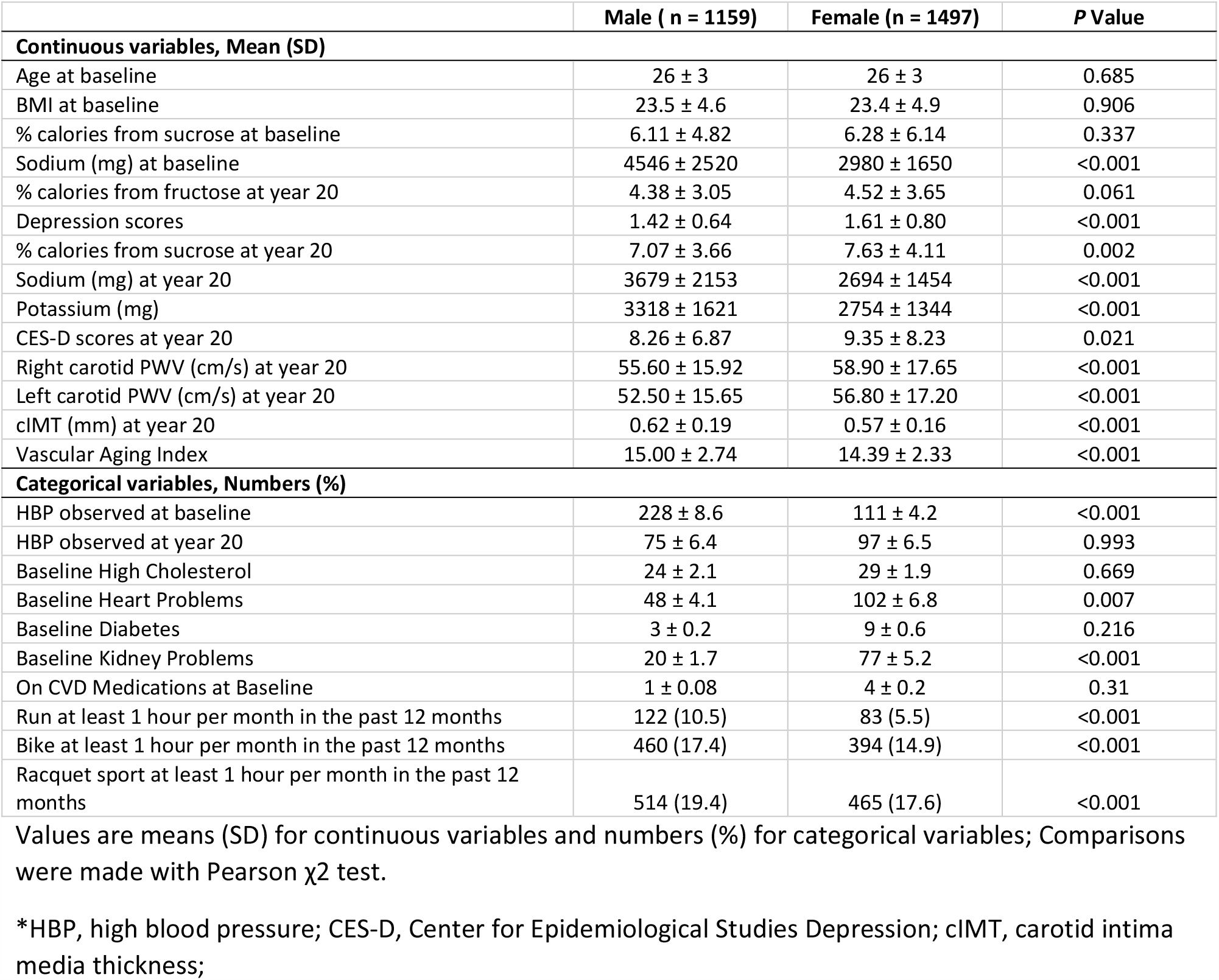
Baseline and year 20 characteristics of the study sample stratified by sex.

|  | Male ( n = 1159) | Female (n = 1497) | P Value |
| --- | --- | --- | --- |
| **Continuous variables, Mean (SD)** |  |  |  |
| Age at baseline | 26 ± 3 | 26 ± 3 | 0.685 |
| BMI at baseline | 23.5 ± 4.6 | 23.4 ± 4.9 | 0.906 |
| % calories from sucrose at baseline | 6.11 ± 4.82 | 6.28 ± 6.14 | 0.337 |
| Sodium (mg) at baseline | 4546 ± 2520 | 2980 ± 1650 | <0.001 |
| % calories from fructose at year 20 | 4.38 ± 3.05 | 4.52 ± 3.65 | 0.061 |
| Depression scores | 1.42 ± 0.64 | 1.61 ± 0.80 | <0.001 |
| % calories from sucrose at year 20 | 7.07 ± 3.66 | 7.63 ± 4.11 | 0.002 |
| Sodium (mg) at year 20 | 3679 ± 2153 | 2694 ± 1454 | <0.001 |
| Potassium (mg) | 3318 ± 1621 | 2754 ± 1344 | <0.001 |
| CES-D scores at year 20 | 8.26 ± 6.87 | 9.35 ± 8.23 | 0.021 |
| Right carotid PWV (cm/s) at year 20 | 55.60 ± 15.92 | 58.90 ± 17.65 | <0.001 |
| Left carotid PWV (cm/s) at year 20 | 52.50 ± 15.65 | 56.80 ± 17.20 | <0.001 |
| cIMT (mm) at year 20 | 0.62 ± 0.19 | 0.57 ± 0.16 | <0.001 |
| Vascular Aging Index | 15.00 ± 2.74 | 14.39 ± 2.33 | <0.001 |
| **Categorical variables, Numbers (%)** |  |  |  |
| HBP observed at baseline | 228 ± 8.6 | 111 ± 4.2 | <0.001 |
| HBP observed at year 20 | 75 ± 6.4 | 97 ± 6.5 | 0.993 |
| Baseline High Cholesterol | 24 ± 2.1 | 29 ± 1.9 | 0.669 |
| Baseline Heart Problems | 48 ± 4.1 | 102 ± 6.8 | 0.007 |
| Baseline Diabetes | 3 ± 0.2 | 9 ± 0.6 | 0.216 |
| Baseline Kidney Problems | 20 ± 1.7 | 77 ± 5.2 | <0.001 |
| On CVD Medications at Baseline | 1 ± 0.08 | 4 ± 0.2 | 0.31 |
| Run at least 1 hour per month in the past 12 months | 122 (10.5) | 83 (5.5) | <0.001 |
| Bike at least 1 hour per month in the past 12 months | 460 (17.4) | 394 (14.9) | <0.001 |
| Racquet sport at least 1 hour per month in the past 12 months | 514 (19.4) | 465 (17.6) | <0.001 |

Values are means (SD) for continuous variables and numbers (%) for categorical variables; Comparisons were made with Pearson $\chi^2$ test.
\*HBP, high blood pressure; CES-D, Center for Epidemiological Studies Depression; cIMT, carotid intima media thickness;

### Association of demographic, dietary and stress predictors with VAI

Dietary, depression and demographic predictors of mid-life VAI are shown in Table 2. We completed multivariate linear regression analyses for factors measured at baseline, when the cohort had a median age of 26 years, as well as when predictors were measured 20 years later, which is the same time point at which carotid ultrasounds were obtained and at which we calculated VAI. The predictors included in the multivariable model were dietary sucrose/fructose, sodium, potassium, age, BMI, race, aerobic exercise and obsHBP. By considering these predictors at baseline, we evaluated their persistent effects on VAI, while by considering them at the time when VAI was obtained we evaluated their acute effects on VAI. For the predictors measured at baseline, age was the only factor significantly associated with increased VAI in both males and females (B-weight = 0.163, p < 0.001, and B-weight=0.154, p < 0.001, respectively), while only in females an association was observed for CES-D scores (B-weight 0.063, p = 0.015). Some sex-specific patterns were observed for the acute effects of the predictors on VAI. In males, sodium intake was directly correlated (B-weight = 0.145, p = 0.003) and potassium intake was inversely correlated with VAI (B -weight = -0.160, p <0.001). Additionally, in males, having had at least 1 hour per month of aerobic exercise (run, bike or racquet sport) consistently over the past 12 months was inversely correlated with VAI (B-weight= -0.85, p = 0.007). In both males and females, age was directly correlated with VAI (B-weight = 0.171, p < 0.001, and B-weight = 0.127, p < 0.001).

**Table 2.** Dietary, depression and demographic predictors of mid-life VAI.

|  | Male |  |  | Female |  |  |
| --- | --- | --- | --- | --- | --- | --- |
|  | b-weight | B-weight | P Value | b-weight | B-weight | P Value |
| **Predictors measured at baseline** |  |  |  |  |  |  |
| Age | **0.130** | **0.163** | **<0.001** | **0.100** | **0.154** | **<0.001** |
| Depression scores | 0.067 | 0.016 | 0.598 | **0.186** | **0.063** | **0.015** |
| Dietary sodium | 0.000 | 0.037 | 0.212 | 0.000 | 0.004 | 0.884 |
| % calories from sucrose | 0.016 | 0.027 | 0.363 | 0.004 | 0.012 | 0.661 |
| BMI | 0.019 | 0.032 | 0.274 | 0.005 | 0.011 | 0.678 |
| obsHBP | 0.345 | 0.050 | 0.094 | -0.039 | -0.004 | 0.866 |
| **Predictors measured at year 20 of follow-up** |  |  |  |  |  |  |
| Age | **0.137** | **0.171** | **<0.001** | **0.079** | **0.127** | **<0.001** |
| CES-D score | -0.008 | -0.021 | 0.514 | 0.009 | 0.033 | 0.236 |
| % calories from fructose at year 20 | -0.028 | -0.031 | 0.349 | 0.026 | 0.042 | 0.157 |
| Dietary sodium | **0.000** | **0.145** | **0.003** | 0.000 | 0.023 | 0.575 |
| Dietary potassium | **0.000** | **-0.160** | **<0.001** | 0.000 | -0.059 | 0.150 |
| obsHBP | 0.110 | 0.010 | 0.757 | 0.101 | 0.011 | 0.694 |
| Aerobic activity (run/bike/racquet sport) | **-0.763** | **-0.085** | **0.007** | -0.400 | -0.042 | 0.137 |

Unstandardized (b-weight) and standardized (B-weight) regression coefficients, significant predictors are bolded for emphasis; n = 1159 (males), n= 1497 (females). ObsHBP, BP > 130/80 mmHg; CES-D, Center for Epidemiological Studies Depression. Collinearity of regression coefficients was ruled out with Pearson correlation and Spearman's rho.

### Predictors of CVD, stroke and death in the CARDIA cohort

To determine whether the dietary predictors, depression and VAI affect cardiovascular morbidity and mortality in the CARDIA cohort, we completed multivariate Cox proportional hazards analysis shown in Table 3. The latest assessment of outcomes in the CARDIA cohort was in 2021, reporting 178 cases (6.7%) of fatal and non-fatal CVD, 61 cases (2.3%) of fatal and non-fatal stroke and 239 (9.0%) all-cause deaths.

**Table 3.**
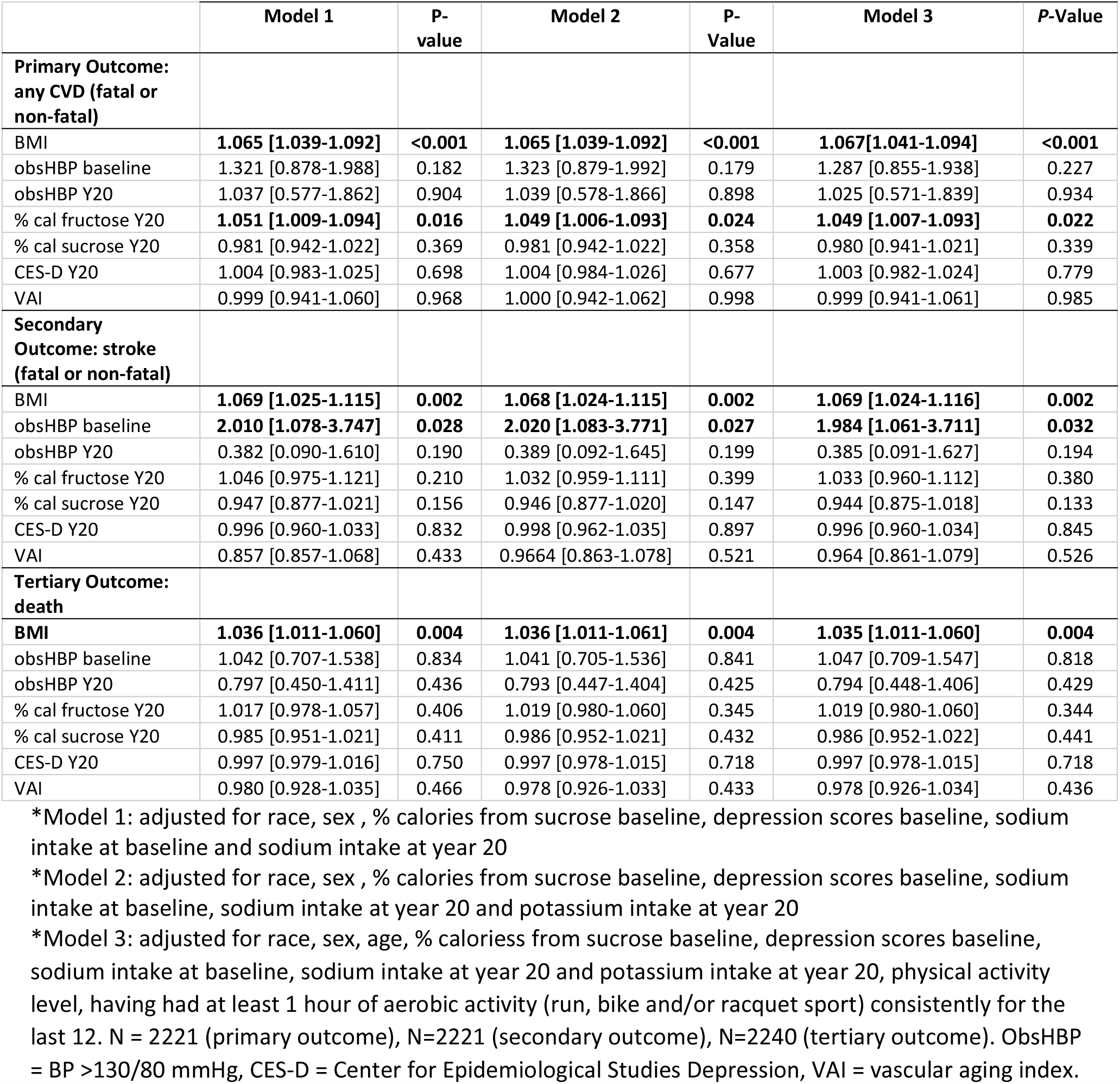
Multivariate Cox proportional hazards analysis: Primary, secondary and tertiary outcomes in all patients with regards to dietary, depression and demographic predictors.

#### PRIMARY OUTCOME: CVD

In Model 1 adjusted for race, sex, age, baseline measures of sucrose intake (expressed as % total calories), CES-D scores, and sodium intake both at baseline and year 20 of follow-up, factors associated with the primary outcome (any CVD fatal and non-fatal) were BMI of participants recorded at baseline (HR = 1.065, 95% CI [1.039-1.092], p <0.001) and percent calories from fructose recorded at year 20 of follow-up (HR = 1.051, 95% CI [1.009-1.094], p = 0.016). Model 2 was additionally adjusted with potassium intake at year 20 of follow-up, and shows the same predictors to be significantly associated with the primary outcome: BMI (HR = 1.067, 95% CI [1.041-1.094], p <0.001) and fructose intake (HR = 1.049, 95% CI [1.006-1.093], p <0.024). In Model 3 we added the covariates of physical activity, both as a self-reported physical activity level as well as specific metrics of consistent aerobic activity. Covariates added to Model 3 included physical activity level, and having had at least 1 hour of aerobic activity (run, bike and/or racquet sport) for 1 hour per month consistently for the last 12 months. Even after the adjustment for physical activity, BMI was still a significant predictor of CVD (HR = 1.041, 95% CI [1.041-1.094], p <0.001) as was dietary fructose (HR = 1.049, 95% CI [1.007-1.093], p <0.022).

#### SECONDARY OUTCOME: STROKE

BMI of the cohort at baseline was also significantly associated with the secondary outcome, stroke (fatal and non-fatal) (HR = 1.069, 95% CI [1.025-1.115], p=0.002), as was having BP above 130/80 mmHg (obsHBP) at the same timepoint (HR = 2.002, 95% CI [1.078-3.747], p = 0.028). Additional covariates in Models 2 and 3 did not change these associations. In Model 2, BMI was associated with HR = 1.068, 95% CI [1.024-1.115], p=0.002 and obsHBP was associated with HR = 2.020, 95% CI [1.083-3.771], p=0.027. In Model 3, the significance of BMI (HR = 1.069, 95% CI [1.024-1.116], p=0.002) and obsHBP (HR = 1.984, 95% CI [1.061-3.711], p=0.032) were maintained.

#### TERTIARY OUTCOME: DEATH

Baseline BMI recorded when the cohort was in their young adulthood was the only predictor significantly associated with all-cause mortality more than three decades later in all three models. In Model 1, HR = 1.036, 95% CI [1.011-1.061], p = 0.004. In Model 2, HR = 1.036, 95% CI [1.011-1.061], p = 0.004. In Model 3, HR = 1.035, 95% CI [1.011-1.060], p = 0.004.

## DISCUSSION

We designed this retrospective observational study to assess how dietary behaviors and psychological stress in young adult White and Black males and females impact vascular aging and CVD risk by mid-life. We found the predictors of VAI at mid-life to be sex-specific whereby dietary sodium, potassium and aerobic activity were significant predictors in males, while depression scores during young adulthood were a significant predictor in females. Moreover, fructose consumption in mid-life was found to be a significant predictor of CVD 15 years later. Likewise, BP > 130/80 mmHg during young adulthood, irrespective of hypertension diagnosis, was found to be associated with an outcome of stroke 35 years later. Lastly, BMI measured during young adulthood was a significant predictor of CVD, stroke, and death.

Our finding that the predictors of vascular aging at mid-life are sex-specific supports the importance of stratifying data by sex, especially data that includes pre-menopausal females. VAI was found to be higher in males compared to females (15.00 ± 2.74 vs. 14.39 ± 2.33, p <0.001, respectively; Table 1). We found that the fructose and sodium intake when the CARDIA cohort was in their adolescent age were not significantly correlated with VAI at mid-life, but that psychological stress was a significant predictor of VAI in females only. Given that depression scores did not differ between males and females (Table 1), this reflects a direct, persistent effect on vascular aging in females only. Similar to other risk factors of vascular aging, psychological stress causes endothelial dysfunction and vascular inflammation ^9^. It is of note that significant associations between VAI and depression scores at year 20 were not observed in our analysis, suggesting that chronic psychological stress, rather than acute, has a negative impact on vascular aging in females. It will be important to investigate the persistent associations between psychological stress and VAI in this aging cohort, especially given the transition to menopause that is occurring concurrently at the time of this writing. When considering acute predictors of VAI, we found that in males only, dietary sodium was directly correlated with VAI (B-weight = 0.145, p = 0.003), while dietary potassium appears to be protective (B-weight = -0.160, p < 0.001). Additionally, having at least 1 hour of aerobic activity (run, bike or racquet sport) was also negatively correlated with VAI (B-weight = - 0.085, p = 0.007) in males. This suggests that in mid-life age, decreasing dietary sodium, while increasing dietary potassium and aerobic activity for males appears to confer protection against vascular aging. It is tempting to speculate that perhaps at least one of the reasons why we did not observe such associations between sodium, potassium and aerobic activity with VAI in females is because their contributions are overpowered by estrogen-dependent β-adrenergic mediated vasodilation ^42^.

In our multivariable Cox regression analyses, only fructose consumption at year 20 and BMI at baseline predicted CVD (primary outcome). Even after we adjusted for potassium intake (Model 2) and consistent aerobic activity (Model 3), these associations still persisted (Table 3). A number of epidemiological studies and subsequent meta-analyses have shown that sugar sweetened beverages increase the risk of hypertension and CVD morbidity and mortality ^35,43^. However, few studies have yet to explore the relative impact of various types of sugars on CVD risk. To our knowledge, this is the first study to demonstrate in a longitudinal cohort that fructose, not sucrose, predicts CVD risk. Here we corroborated the preclinical data which demonstrated that dietary fructose contributes to vascular stiffness both in adolescence and adulthood ^29, 30, 33^. In addition to vascular changes, high fructose consumption has been shown to reduce plasma insulin and leptin levels and increase ghrelin concentrations ^44^, which may contribute to obesity and thus a pro-inflammatory state. While our findings suggest that fructose is an independent predictor of CVD, it may also contribute to obesity, which is agreeable with our finding that BMI is another significant predictor of CVD morbidity in the CARDIA cohort. In our analysis for the secondary outcome, stroke, we found that both BMI and having BP > 130/80 mmHg, in adolescence were significant predictors in all three models. We chose to assess actual measurements of BP, independent of hypertension diagnosis, because we believe that this is a more clinically relevant scenario contributing to vascular aging. Persistent shear stress caused by elevated BP is known to cause local and functional changes to vascular compliance ^45^. This highlights the importance of BP control, and the elevated risk in scenarios when such is not achieved even with medical therapy. Here we show that BP > 130/80 mmHg in adolescence doubles the risk of having a stroke 35 years later. BMI in adolescence was the only variable in our study to predict all three outcomes: CVD, stroke, and death. The link between obesity and CVD has been shown to be multi-faceted and includes interactions between environment, socioeconomic status, genetics, physical activity behaviors, and internal factors ^46^. A prolonged state of energy imbalance (greater energy intake compared to expenditure) leads to changes in the functions of adipose tissue, including increased pro-inflammatory adipokine production. The pro-inflammatory state causes atherogenesis and increased endothelial vasomotor tone, both of which can contribute to CVD risk ^47^.

A limitation of the present study is its retrospective observational nature. Furthermore, dietary practices are derived from Diet History Questionnaires, which are associated with inherent social desirability bias. Compared to other dietary history methods (i.e. 24-hour recall or food records), the diet history method used in the CARDIA study may overestimate absolute intake of nutrients, but this should not interfere with analyses that employ comparisons of means among different subgroups within the CARDIA study and only becomes a potential problem when comparing intakes to populations outside of the CARDIA cohort. Every effort was made to keep the diet history questionnaire at all visits consistent. Regarding outliers and missing values, the Nutrition Working Group during visit 1 decided to include all out-of-range values in the participants’ records and to not impute any missing data on serving amounts or frequency.

## CONCLUSION

In conclusion, our findings suggest that dietary and lifestyle practices that impact vascular aging during adolescence and into mid-life are sex-specific. Lowering psychological stress was found to be important in females, while lowering dietary sodium, and increasing potassium and aerobic activity was found to be significantly correlated with vascular aging in males. Furthermore, limiting fructose consumption may confer cardiovascular protection later into mid-life. High fructose consumption may contribute to a higher BMI, which we found to be a significant predictor of CVD, stroke, and death. Given the aging population in the U.S. and the challenges of large-scale behavior change, there is a need to find small-scale behavior changes that can help prevent the vascular aging process^10,48^. This study provides evidence that focus on nutrient-specific changes (i.e. lowering fructose in males and females, and focus on sodium and potassium in males) that are within the purview of overall dietary pattern recommendations, may at least partially mitigate vascular aging in early adulthood and the consequential CVD risk in mid-life.

## Data Availability

The data is available upon reasonable request from the corresponding author.

## Abbreviations

aPWV: aortic pulse-wave velocity
CARDIA: Coronary Artery Risk Development in Young Adults Study
cfPWV: carotid-femoral pulse wave velocity
cIMT: carotid intima–media thickness
CVD: cardiovascular diseases
HFCS: high fructose corn syrup
NO: nitric oxide
obsHBP: Observed high blood pressure (BP > 130/80 mmHg)
VAI: vascular aging index

## DATA AVAILABILITY

This study was conducted using the Coronary Artery Risk Development In young Adults (CARDIA) research materials obtained from the National Heart, Lung, and Blood Institute (NIH-NHLBI) Biologic Specimen and Data Repository Information Coordinating Center. Individuals wishing to obtain data from the CARDIA study must do so by entering in the NHLBI Research Materials Distribution Agreement (RMDA). This agreement has been most recently completed by the corresponding author (DK) on 4/15/2022.

## GRANTS

None.

## DISCLOSURES

The authors have no conflict of interest.

## DISCLAIMERS

None.

## AUTHOR CONTRIBUTIONS

Conceived and designed research (DK), analyzed data (MO, AB, DP, EF, AV), interpreted results of experiments (DK), prepared figures (DK, MO), drafted manuscript (MO, DK), edited and revised manuscript (MO, EF, AB, AV, DP, DK), approved final version of manuscript (MO, EF, AB, AV, DP, DK).

## Notes

### Competing Interest Statement

The authors have declared no competing interest.

### Author Declarations

IRB# 063319MP2X, Wayne State University, Detroit, MI

## REFERENCES

1. Tsao CW, Aday AW, Almarzooq ZI, Alonso A, Beaton AZ, Bittencourt MS, Boehme AK, Buxton AE, Carson AP, Commodore-Mensah Y, Elkind MSV, Evenson KR, Eze-Nliam C, Ferguson JF, Generoso G, Ho JE, Kalani R, Khan SS, Kissela BM, Knutson KL, Levine DA, Lewis TT, Liu J, Loop MS, Ma J, Mussolino ME, Navaneethan SD, Perak AM, Poudel R, Rezk-Hanna M, Roth GA, Schroeder EB, Shah SH, Thacker EL, VanWagner LB, Virani SS, Voecks JH, Wang NY, Yaffe K and Martin SS. Heart Disease and Stroke Statistics-2022 Update: A Report From the American Heart Association. Circulation. 2022;145:e153–e639.

2. Mikael LR, Paiva AMG, Gomes MM, Sousa ALL, Jardim P, Vitorino PVO, Euzebio MB, Sousa WM and Barroso WKS. Vascular Aging and Arterial Stiffness. Arq Bras Cardiol. 2017;109:253–258.

3. Zhong Q, Hu MJ, Cui YJ, Liang L, Zhou MM, Yang YW and Huang F. Carotid-Femoral Pulse Wave Velocity in the Prediction of Cardiovascular Events and Mortality: An Updated Systematic Review and Meta-Analysis. Angiology. 2018;69:617–629.

4. Nilsson Wadstrom B, Fatehali AH, Engstrom G and Nilsson PM. A Vascular Aging Index as Independent Predictor of Cardiovascular Events and Total Mortality in an Elderly Urban Population. Angiology. 2019;70:929–937.

5. Ungvari Z, Tarantini S, Donato AJ, Galvan V and Csiszar A. Mechanisms of Vascular Aging. Circ Res. 2018;123:849–867.

6. Harvey A, Montezano AC, Lopes RA, Rios F and Touyz RM. Vascular Fibrosis in Aging and Hypertension: Molecular Mechanisms and Clinical Implications. Can J Cardiol. 2016;32:659–68.

7. LaRocca TJ, Martens CR and Seals DR. Nutrition and other lifestyle influences on arterial aging. Ageing Res Rev. 2017;39:106–119.

8. Merz AA and Cheng S. Sex differences in cardiovascular ageing. Heart. 2016;102:825–31.

9. Sara JDS, Toya T, Ahmad A, Clark MM, Gilliam WP, Lerman LO and Lerman A. Mental Stress and Its Effects on Vascular Health. Mayo Clin Proc. 2022;97:951–990.

10. Vespa J, Armstrong DM and Medina L. Demographic turning points for the United States: Population projections for 2020 to 2060: US Department of Commerce, Economics and Statistics Administration, US …; 2018.

11. Sabbatini AR and Kararigas G. Menopause-Related Estrogen Decrease and the Pathogenesis of HFpEF: JACC Review Topic of the Week. J Am Coll Cardiol. 2020;75:1074–1082.

12. Stanhewicz AE, Wenner MM and Stachenfeld NS. Sex differences in endothelial function important to vascular health and overall cardiovascular disease risk across the lifespan. Am J Physiol Heart Circ Physiol. 2018;315:H1569–H1588.

13. Moreau KL. Modulatory influence of sex hormones on vascular aging. Am J Physiol Heart Circ Physiol. 2019;316:H522–H526.

14. Ji H, Kwan AC, Chen MT, Ouyang D, Ebinger JE, Bell SP, Niiranen TJ, Bello NA and Cheng S. Sex Differences in Myocardial and Vascular Aging. Circ Res. 2022;130:566–577.

15. Kuehner C. Why is depression more common among women than among men? Lancet Psychiatry. 2017;4:146–158.

16. Hankin BL, Mermelstein R and Roesch L. Sex differences in adolescent depression: stress exposure and reactivity models. Child Dev. 2007;78:279–95.

17. Plaisier I, de Bruijn JG, Smit JH, de Graaf R, Ten Have M, Beekman AT, van Dyck R and Penninx BW. Work and family roles and the association with depressive and anxiety disorders: differences between men and women. J Affect Disord. 2008;105:63–72.

18. Connelly PJ, Azizi Z, Alipour P, Delles C, Pilote L and Raparelli V. The Importance of Gender to Understand Sex Differences in Cardiovascular Disease. Can J Cardiol. 2021;37:699–710.

19. Lloyd-Jones DM, Allen NB, Anderson CAM, Black T, Brewer LC, Foraker RE, Grandner MA, Lavretsky H, Perak AM, Sharma G, Rosamond W and American Heart A. Life’s Essential 8: Updating and Enhancing the American Heart Association’s Construct of Cardiovascular Health: A Presidential Advisory From the American Heart Association. Circulation. 2022;146:e18–e43.

20. Rossman MJ, LaRocca TJ, Martens CR and Seals DR. Healthy lifestyle-based approaches for successful vascular aging. J Appl Physiol (1985). 2018;125:1888–1900.

21. Nowak KL, Rossman MJ, Chonchol M and Seals DR. Strategies for Achieving Healthy Vascular Aging. Hypertension. 2018;71:389–402.

22. D’Elia L, Galletti F, La Fata E, Sabino P and Strazzullo P. Effect of dietary sodium restriction on arterial stiffness: systematic review and meta-analysis of the randomized controlled trials. J Hypertens. 2018;36:734–743.

23. Ezekowitz JA, Colin-Ramirez E, Ross H, Escobedo J, Macdonald P, Troughton R, Saldarriaga C, Alemayehu W, McAlister FA, Arcand J, Atherton J, Doughty R, Gupta M, Howlett J, Jaffer S, Lavoie A, Lund M, Marwick T, McKelvie R, Moe G, Pandey AS, Porepa L, Rajda M, Rheault H, Singh J, Toma M, Virani S, Zieroth S and Investigators S-H. Reduction of dietary sodium to less than 100 mmol in heart failure (SODIUM-HF): an international, open-label, randomised, controlled trial. Lancet. 2022;399:1391–1400.

24. McDonough AA, Veiras LC, Guevara CA and Ralph DL. Cardiovascular benefits associated with higher dietary K(+) vs. lower dietary Na(+): evidence from population and mechanistic studies. Am J Physiol Endocrinol Metab. 2017;312:E348–E356.

25. Houston MC. The importance of potassium in managing hypertension. Curr Hypertens Rep. 2011;13:309–17.

26. Walker RW, Dumke KA and Goran MI. Fructose content in popular beverages made with and without high-fructose corn syrup. Nutrition. 2014;30:928–935.

27. Ventura EE, Davis JN and Goran MI. Sugar content of popular sweetened beverages based on objective laboratory analysis: focus on fructose content. Obesity (Silver Spring). 2011;19:868–74.

28. Tappy L and Le KA. Metabolic effects of fructose and the worldwide increase in obesity. Physiol Rev. 2010;90:23–46.

29. Komnenov D, Levanovich PE, Perecki N, Chung CS and Rossi NF. Aortic Stiffness and Diastolic Dysfunction in Sprague Dawley Rats Consuming Short-Term Fructose Plus High Salt Diet. Integr Blood Press Control. 2020;13:111–124.

30. Komnenov D and Rossi NF. Fructose-induced salt-sensitive blood pressure differentially affects sympathetically mediated aortic stiffness in male and female Sprague-Dawley rats. Physiol Rep. 2023;11:e15687.

31. Komnenov D, Levanovich PE and Rossi NF. Hypertension Associated with Fructose and High Salt: Renal and Sympathetic Mechanisms. Nutrients. 2019;11.

32. Soncrant T, Komnenov D, Beierwaltes WH, Chen H, Wu M and Rossi NF. Bilateral renal cryodenervation decreases arterial pressure and improves insulin sensitivity in fructose-fed Sprague-Dawley rats. Am J Physiol Regul Integr Comp Physiol. 2018;315:R529–R538.

33. Levanovich PE, Chung CS, Komnenov D and Rossi NF. Fructose plus High-Salt Diet in Early Life Results in Salt-Sensitive Cardiovascular Changes in Mature Male Sprague Dawley Rats. Nutrients. 2021;13.

34. Levanovich PE, Daugherty AM, Komnenov D and Rossi NF. Dietary fructose and high salt in young male Sprague Dawley rats induces salt-sensitive changes in renal function in later life. Physiol Rep. 2022;10:e15456.

35. Jayalath VH, de Souza RJ, Ha V, Mirrahimi A, Blanco-Mejia S, Di Buono M, Jenkins AL, Leiter LA, Wolever TM, Beyene J, Kendall CW, Jenkins DJ and Sievenpiper JL. Sugar-sweetened beverage consumption and incident hypertension: a systematic review and meta-analysis of prospective cohorts. Am J Clin Nutr. 2015;102:914–21.

36. Gao JW, Hao QY, Zhang HF, Li XZ, Yuan ZM, Guo Y, Wang JF, Zhang SL and Liu PM. Low-Carbohydrate Diet Score and Coronary Artery Calcium Progression: Results From the CARDIA Study. Arterioscler Thromb Vasc Biol. 2021;41:491–500.

37. Duffey KJ, Gordon-Larsen P, Steffen LM, Jacobs DR, Jr. and Popkin BM. Drinking caloric beverages increases the risk of adverse cardiometabolic outcomes in the Coronary Artery Risk Development in Young Adults (CARDIA) Study. Am J Clin Nutr. 2010;92:954–9.

38. Friedman GD, Cutter GR, Donahue RP, Hughes GH, Hulley SB, Jacobs Jr DR, Liu K and Savage PJ. CARDIA: study design, recruitment, and some characteristics of the examined subjects. Journal of clinical epidemiology. 1988;41:1105–1116.

39. Jacobs DR, Jr., Yatsuya H, Hearst MO, Thyagarajan B, Kalhan R, Rosenberg S, Smith LJ, Barr RG and Duprez DA. Rate of decline of forced vital capacity predicts future arterial hypertension: the Coronary Artery Risk Development in Young Adults Study. Hypertension. 2012;59:219–25.

40. Whelton PK and Carey RM. The 2017 American College of Cardiology/American Heart Association clinical practice guideline for high blood pressure in adults. JAMA cardiology. 2018;3:352–353.

41. Jacobs DR, Jr., Hahn LP, Haskell WL, Pirie P and Sidney S. Validity and Reliability of Short Physical Activity History: Cardia and the Minnesota Heart Health Program. J Cardiopulm Rehabil. 1989;9:448–459.

42. Baker SE, Limberg JK, Ranadive SM and Joyner MJ. Neurovascular control of blood pressure is influenced by aging, sex, and sex hormones. Am J Physiol-Reg I. 2016;311:R1271–R1275.

43. Yin J, Zhu Y, Malik V, Li X, Peng X, Zhang FF, Shan Z and Liu L. Intake of Sugar-Sweetened and Low-Calorie Sweetened Beverages and Risk of Cardiovascular Disease: A Meta-Analysis and Systematic Review. Adv Nutr. 2021;12:89–101.

44. Stanhope KL, Schwarz JM, Keim NL, Griffen SC, Bremer AA, Graham JL, Hatcher B, Cox CL, Dyachenko A, Zhang W, McGahan JP, Seibert A, Krauss RM, Chiu S, Schaefer EJ, Ai M, Otokozawa S, Nakajima K, Nakano T, Beysen C, Hellerstein MK, Berglund L and Havel PJ. Consuming fructose-sweetened, not glucose-sweetened, beverages increases visceral adiposity and lipids and decreases insulin sensitivity in overweight/obese humans. J Clin Invest. 2009;119:1322–34.

45. Siasos G, Tsigkou V, Coskun AU, Oikonomou E, Zaromitidou M, Lerman LO, Lerman A and Stone PH. The Role of Shear Stress in Coronary Artery Disease. Curr Top Med Chem. 2023.

46. Pigeyre M, Yazdi FT, Kaur Y and Meyre D. Recent progress in genetics, epigenetics and metagenomics unveils the pathophysiology of human obesity. Clin Sci (Lond). 2016;130:943–86.

47. Redinger RN. The pathophysiology of obesity and its clinical manifestations. Gastroenterology & hepatology. 2007;3:856.

48. Conner M and Norman P. Health behaviour: Current issues and challenges. Psychol Health. 2017;32:895–906.

